# Sleep disorders as risk factors for calcific aortic stenosis

**DOI:** 10.1101/2024.11.15.24317390

**Authors:** Nadim El Jamal, Thomas G. Brooks, Carsten Skarke, Garret A. FitzGerald

## Abstract

**Background:** Circadian disruption and sleep disorders have been shown to increase the risk for many cardiovascular diseases. Their association specifically with valvular heart disease, however, is inconclusive.

**Methods:** We leveraged large electronic health record datasets (the TriNetX network and the All *of* Us study) to test whether sleep disorders are a risk factor for calcific aortic stenosis (AS). We fitted Cox proportional hazards models to quantify the risk of future incidence of AS in patients with sleep disorders. We also explored clinical laboratory test datasets for biochemical signals that might explain the association, running mediation analyses.

**Results:** In our fully adjusted Cox models, we find that having any sleep disorder increases the risk for the future incidence of AS (HR: 1.15 95% CI: 1.13-1.18). Changes in lipid profile mediate a proportion of that association.

**Conclusion:** Sleep disorders are associated with an increased risk of AS incidence. That association is independent of classical cardiovascular risk factors even though dyslipidemia plays a large role in mediating this risk.

## Introduction

Modern lifestyles are full of disruptors of our normal sleep-wake rhythms, such as artificial light, noise, trans meridian travel, as well as social and professional demands. Sleep disturbances show high prevalence, for example, 36.8% of respondents to a CDC survey reported a short sleep duration (less than 7 hours) in 2022.^1^ A clear signal emerged from epidemiological studies linking disrupted sleep-wake rhythms (measured by sleep time, sleep duration, or working as a shift-worker) to an increased risk for cardiovascular disease (CVD).^2–4^ Sleep disorders (diagnosed and self-reported) have also been linked to an increased risk of various cardiovascular diseases.^5–7^ These studies do not include valvular disease as a clinical outcome measure. To date, only one study from the UK-biobank tests the association between disrupted sleep wake rhythms and aortic stenosis (AS) and concludes that self-reported insomnia is linked to the development of AS.^8^

AS is strongly associated with aging and is estimated to be prevalent in 5-10% of adults older than 65 years; its incidence is predicted to increase along with prolonged life expectancy.^9^ AS has an overlapping risk factor profile with atherosclerosis which highlights a shared pathogenic processes with the initiation phase of AS. Circadian disruption and sleep disorders have a well-established link to higher risks of coronary artery disease.^2, 3, 7^ We thus hypothesized that circadian disruption caused by sleep disorders increases the risk for AS development.

In this study we test the association between sleep disorders and the future incidence of aortic stenosis using two large electronic health record (EHR) databases. We also leverage biochemical data for potential mechanistic insights into that association.

## Methods

### Patients

The data used in this study were downloaded on April 27, 2023, from the TriNetX Diamond Network which provided access to electronic medical records from approximately 108 million patients across 92 healthcare organizations. For our testing cohort, we obtained EHR data from 8.8 million patients above age 50, with a measurement of height and weight, and with a diagnosis of sleep disorders (ICD10 code G47, ICD9 code 327) and from a random sample of 8.8 million patients who meet the same criteria but without a diagnosis of sleep disorders.

Our study was deemed exempt from IRB review by the institutional review board at the University of Pennsylvania (Protocol # 853440).

For our validation cohort we used data from the All *of* Us study (data version 7) containing EHR data from 392,259 participants 70,070 of whom have been diagnosed with a sleep disorder.

Further exclusion criteria applied to both data sources were hidden sex, a diagnosis of congenital aortic valve disease, a non-physiologic BMI (<12 or >50), the presence of a procedural code for any aortic valve procedure without the presence of an AS diagnosis, and a diagnosis of AS before age 66. All diagnostic, procedural, or prescription codes used in this study are listed in table S1.

### Statistical Analysis

For both the TriNetX and the All *of* US data, we divided the study population into two groups, those with and without a diagnosis of sleep disorders by age 65. Patients who developed sleep disorders after age 65 were included in the group of patients without sleep disorders. We chose the cutoff at 65 years based on previous work on AS risk factors and on its strong association with aging.^10^ To test whether sleep disorders qualify as risk factors for AS, we fitted Cox proportional hazards models comparing patients who were diagnosed with sleep disorders by age 65 to those who were not. The outcome of the model was specified as a diagnosis of non-rheumatic aortic valve stenosis. We used each patient’s age as the time component, and data was censored at patient’s death or the year 2020. We started the follow up time at age 66 to separate the exposure from the outcome by a year and decrease the risk for reverse causation. We ended the follow up period at age 80 (Table 1). We included hypertension, hyperlipidemia, chronic kidney disease, BMI, diabetes, and sex at birth as covariates in the models. We applied the same cutoff of 65 for covariate diagnoses. We tested the proportional hazards assumption by plotting Schoenfeld residuals.^11^ Covariates where the assumption was violated were included as time varying coefficients. When analyzing the incidence of any sleep disorder, the earliest date of any sleep diagnosis was considered the date of sleep disorder incidence in patients with more than one sleep disorder diagnosis.

**Table 1.**
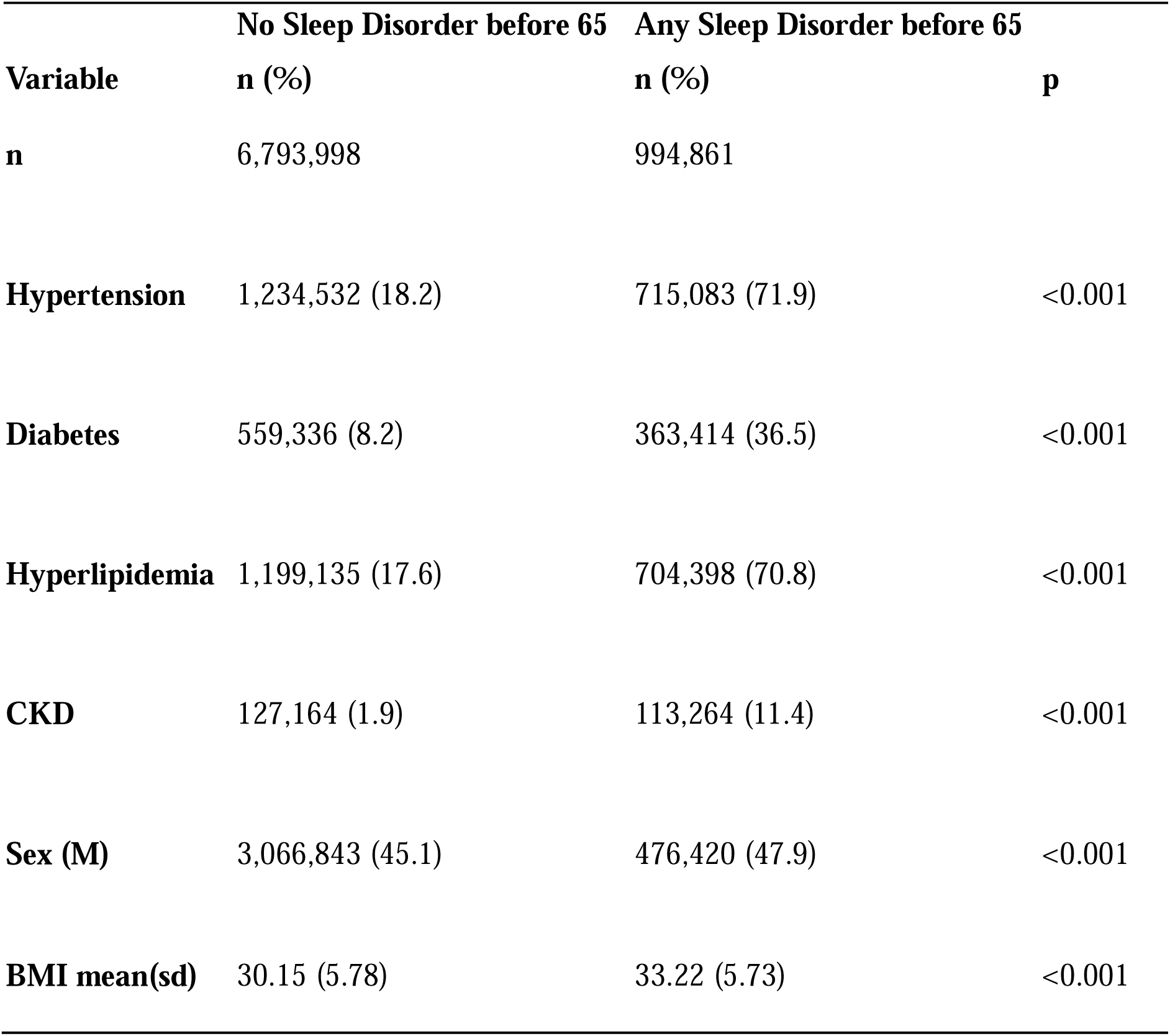
Characteristics of the two patient populations in the TriNetX network from data collected before age 65.

| **Variable** | **No Sleep Disorder before 65** | **Any Sleep Disorder before 65** | **p** |
| --- | --- | --- | --- |
|  | **n (%)** | **n (%)** |  |
| **n** | 6,793,998 | 994,861 |  |
| **Hypertension** | 1,234,532 (18.2) | 715,083 (71.9) | <0.001 |
| **Diabetes** | 559,336 (8.2) | 363,414 (36.5) | <0.001 |
| **Hyperlipidemia** | 1,199,135 (17.6) | 704,398 (70.8) | <0.001 |
| **CKD** | 127,164 (1.9) | 113,264 (11.4) | <0.001 |
| **Sex (M)** | 3,066,843 (45.1) | 476,420 (47.9) | <0.001 |
| **BMI mean(sd)** | 30.15 (5.78) | 33.22 (5.73) | <0.001 |

We used the mediation package in R to quantify the degree to which the above covariates confounded the association between sleep disorders and AS.^12^ We used a survival regression model quantifying the association between the covariate and the development of AS after age 66 and a logistic regression model quantifying the association between having a sleep disorder and the covariate of interest for the mediation analysis. We used our adjusted Cox proportional hazards model to calculate the population attributable fraction (PAF) for all covariates in the model using the graphPAF package in R to quantify further the impact of each covariate on the development of AS,.^13^ PAF describes the fraction of disease cases attributable to a specific exposure.^14^

To explore biochemical signals potentially explicative of the association between sleep disorders and AS, we divided the patients in the TriNetX data into 4 cohorts: (i) those with a sleep disorder diagnosis before age 65 and incident AS after age 66, (ii) those with no prior sleep disorder but incident AS, (iii) those with prior sleep disorders but no incident AS, and (iv) heathy patients with neither prior sleep disorder nor incident AS. We then examined all clinical lab tests in the database performed prior to age 65 and eliminated any outlier values (beyond 1.5 times the interquartile range from the first and second quartiles), and excluded tests not represented across all 4 cohorts. Measurements after age 65 were also eliminated. Multiple test values prior to age 65 for a specific lab test in a single patient were averaged. We then ran a linear regression analysis with the lab test numeric value as the dependent variable and the cohort as the independent variable such that all cohorts are compared to the sleep disorder and incident AS cohort. We selected the lab tests where the Benjamini-Hochberg adjusted *p*-value (*q*-value) was <0.1 and the fold change in the lab numeric value was ≥ 5% for all the three comparisons to make sure the resulting labs are not selected based solely on a low *p*-value due to a large sample size. To further quantify the effect each of the statistically significant differences in lab results have on the association between sleep disorders and AS, we repeated the above-described mediation analysis while replacing our covariates of interest with the labs surviving our *q*-value and fold-change cutoff.

Data management and all analyses were performed using R versions (4.3 and 4.4).

## Results

Multiple sleep disorders are associated with an increased risk for AS incidence.

Figure S1 shows how we arrived at our population of interest from the TriNetX data. In the TriNetX database, patients with sleep disorders had a significantly higher prevalence of cardiovascular risk factors and a significantly higher BMI (Table 1). 10.6% of the studied sample from TriNetX presented with a sleep disorder diagnosis before age 65. The mean age of diagnosis was 62.7±2.4 years with sleep apnea as the most prevalent diagnosis (7.1%, Table S2). There were 133,419 cases of incident AS between ages 66 and 80. Our unadjusted Cox proportional hazards models showed an increased risk of AS incidence with any sleep disorder (HR: 2.61, 95% CI: 2.55-2.67) which remained significant when each individual sleep disorder was interrogated (Figure 1 A). Here, sleep apnea showed the strongest signal (HR: 3.04, 95% CI: 2.96-3.12). The significant risk association between AS incidence and any sleep disorder and sleep apnea survived our adjusted model (HR: 1.15 95% CI: 1.13-1.83 and HR: 1.20 95% CI: 1.17-1.24, respectively) where we controlled for various cardiovascular risk factors. Additional candidate sleep disorders linked to AS incidence included hypersomnia (HR: 1.18, 95%CI: 1.07-1.31), narcolepsy (HR: 1.28 95% CI:1.03-1.60) and sleep movement disorders (HR: 1.19 95% CI: 1.07-1.33, Figure 1 B). Results from the mediation analysis shows significant and negative average causal mediation effects (ACME) and average direct effects (ADE) indicating that the presence of a sleep disorder is significantly associated with smaller AS free survival time independent from the tested covariates, but that these covariates mediate a proportion of that association (Table 2).

**Figure 1.**
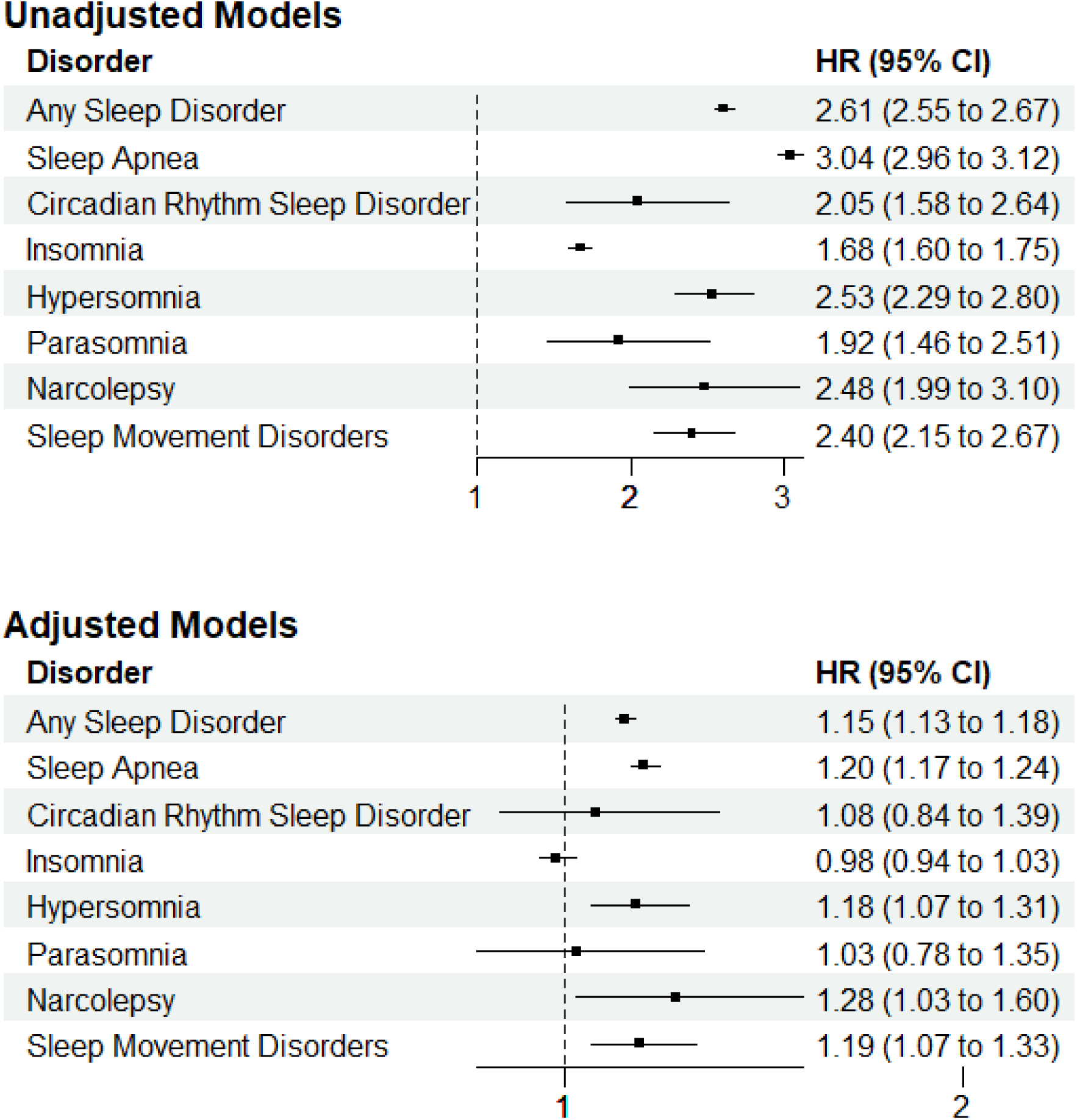
Hazard ratios from unadjusted cox proportional hazards models (Top) and adjusted models (Bottom) testing the association between sleep disorders and incident aortic stenosis in the TriNetX database. Adjusted for hypertension, hyperlipidemia, diabetes, chronic kidney disease, sex at birth, and BMI.

**Table 2.** Results from the mediation analysis quantifying the effects cardiovascular risk factors have on the association between sleep disorders and incident aortic stenosis. Data from the TriNetX database. **ACME:** Average causal mediation effects of sleep disorders on aortic stenosis through the risk factor **ADE:** Average direct effect of sleep disorders on aortic stenosis. **CKD:** chronic kidney disease

| Risk Factor | ACME |  | ADE |  |
| --- | --- | --- | --- | --- |
|  | Estimate | P-value | Estimate | P-value |
| Hypertension | -14.77 | <2e-16 | -15.26 | <2e-16 |
| Hyperlipidemia | -13.10 | <2e-16 | -18.05 | <2e-16 |
| Diabetes | -8.68 | <2e-16 | -21.53 | <2e-16 |
| CKD | -3.08 | <2e-16 | -31.52 | <2e-16 |
| BMI | -7.58 | <2e-16 | -30.62 | <2e-16 |

While sleep disorders are attributed to a smaller proportion of AS cases developed during the follow up period (PAF: 4.35%, 95% CI, 3.75-4.82) compared to other cardiovascular risk factors such as a BMI ≥ 30 (PAF: 33.2%, 95% CI, 32.7-33.9), they still inflict a similar burden to chronic kidney disease (PAF 4.4%, 95% CI: 4.2 -4.67) in the population forming the TriNetX database (Table 3).

**Table 3:** Population Attributable Fraction (PAF) of sleep disorders and the different cardiovascular risk factors.

| **Risk Factor** | **PAF (%)** | **95% CI** |
| --- | --- | --- |
| Sleep Disorders | 4.35 | 3.75-4.82 |
| Chronic Kidney Disease | 4.4 | 4.2-4.67 |
| Hyperlipidemia | 7.52 | 6.6-8.32 |
| Diabetes | 11.9 | 11.5-12.4 |
| Hypertension | 18.8 | 18.0-19.6 |
| BMI ≥ 30 | 33.2 | 32.7-33.9 |

In the All *of* Us cohort, 957 cases of incident AS were present after age 66 The adjusted Cox proportional hazards model confirmed the increased risk for AS development in patients diagnosed with any sleep disorder (HR: 1.40, 95% CI: 1.12-1.77 in the fully adjusted model). Due to the small number of events for several sleep disorders, we did not expand the modeling across each of the sleep disorder categories. We did not conduct a laboratory test analysis due to sample size limitations for each individual test. Figure S2 shows how we arrived at our population of interest from the All *of* Us study and Table S3 shows the characteristics of the patient population from data collected before age 65. As we observed in the TriNetX database, the All *of* Us cohort showed a similar trend of a higher prevalence of cardiovascular risk factors among patients with sleep disorders. All *of* US study patients had a similar sleep disorder prevalence with slightly younger diagnosis ages compared to the TriNetX network (Table S4).

Taken together, patients with sleep disorders are at an increased risk of developing aortic stenosis. This association is independent from cardiovascular risk factors that mediate a proportion of this association.

Changes in lipid metabolism play a role in the increased risk.

In the TriNetX database, 608 lab tests were included in our biochemical analyses. Four lab tests, namely HDL, LDL, total cholesterol, and triglycerides, survived the multiple testing correction at a *q*-value ≤ 0.1 while satisfying a fold change ≥ 5% in all three comparisons with the disease group (AS incidence after age 66 and sleep disorder before age 65, Table S5). Figure 2 shows the distributions of the numeric value of each of the four lab tests across each of the four cohorts.

**Figure 2.**
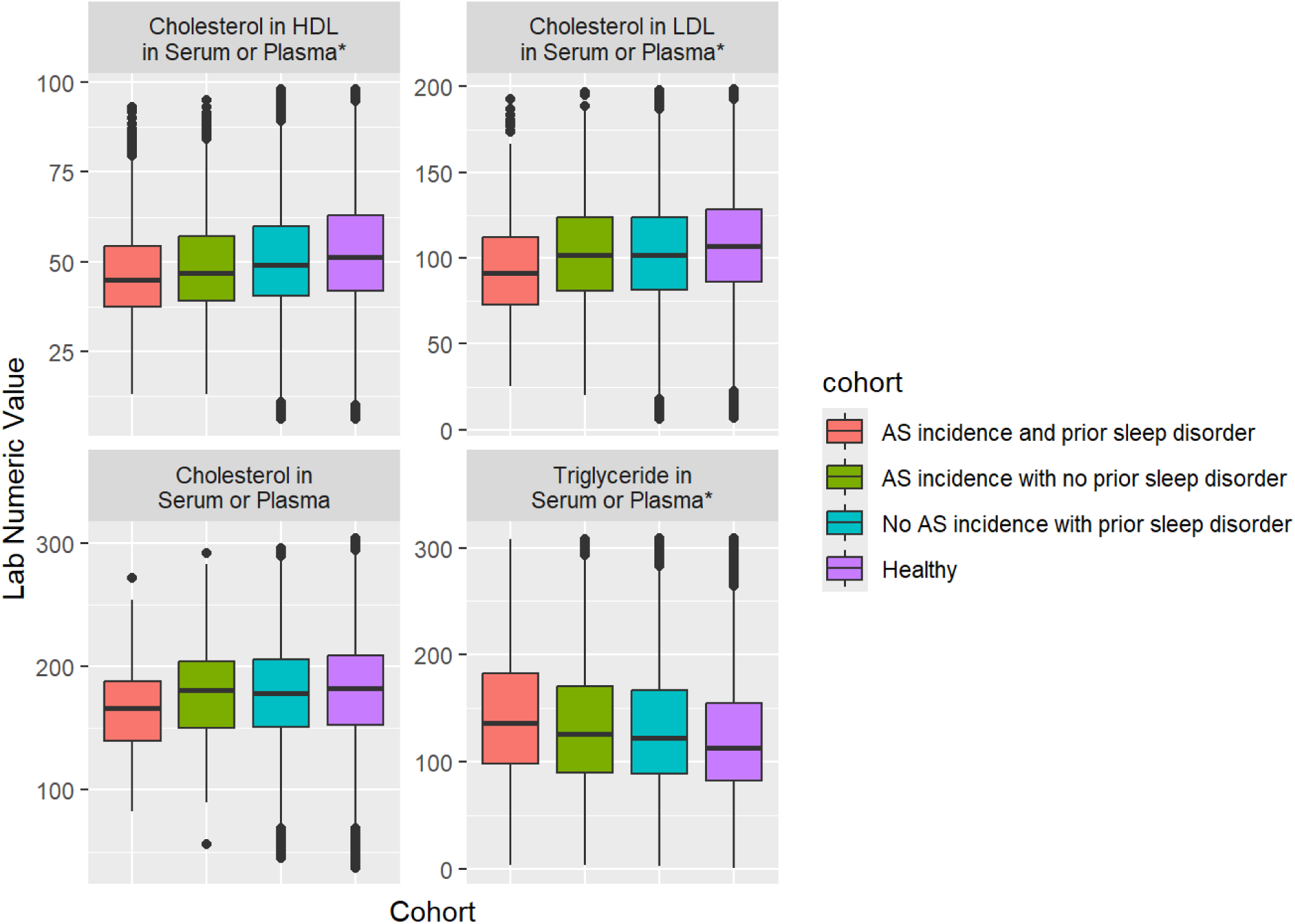
Distribution of lab test numeric values with q-values ≤ 0.1 and fold change ≥5% for all cohort comparisons with the healthy cohort. *q-value ≤ 0.05. Based on data from the TriNetX database.

The distribution of HDL cholesterol was lower and that of triglycerides was higher in the AS and sleep cohort, indicating less favorable lipid profiles (Table S6). LDL and total cholesterol were lower in the diseased cohort. This observation might be driven by statin use which, while effective in lowering LDL, does not seem to decrease AS progression.^15, 16^ Indeed, 77% of patients in the AS incidence and prior sleep disorder cohort were on statins before age 65, a significantly higher proportion than the other cohorts (Chi-squared p value <0.001, Figure S3).

For HDL, LDL, total cholesterol and triglycerides, the mediation analysis showed significant and negative average direct effects (ADE, *p-*values <0.02 for all) indicating that sleep disorders are associated with a shorter AS free survival time independent from these lipid differences. The analysis also showed negative and significant causal mediation effects (ACME, *p-values* <0.01 for all) indicating that a proportion of this association is mediated by changes in these lipid parameters (Table 4). In addition to the fully adjusted Cox proportional hazards models, both covariate and laboratory test mediation analyses support the conclusion that sleep disorders are independently associated with an increased risk for AS incidence.

**Table 4.**
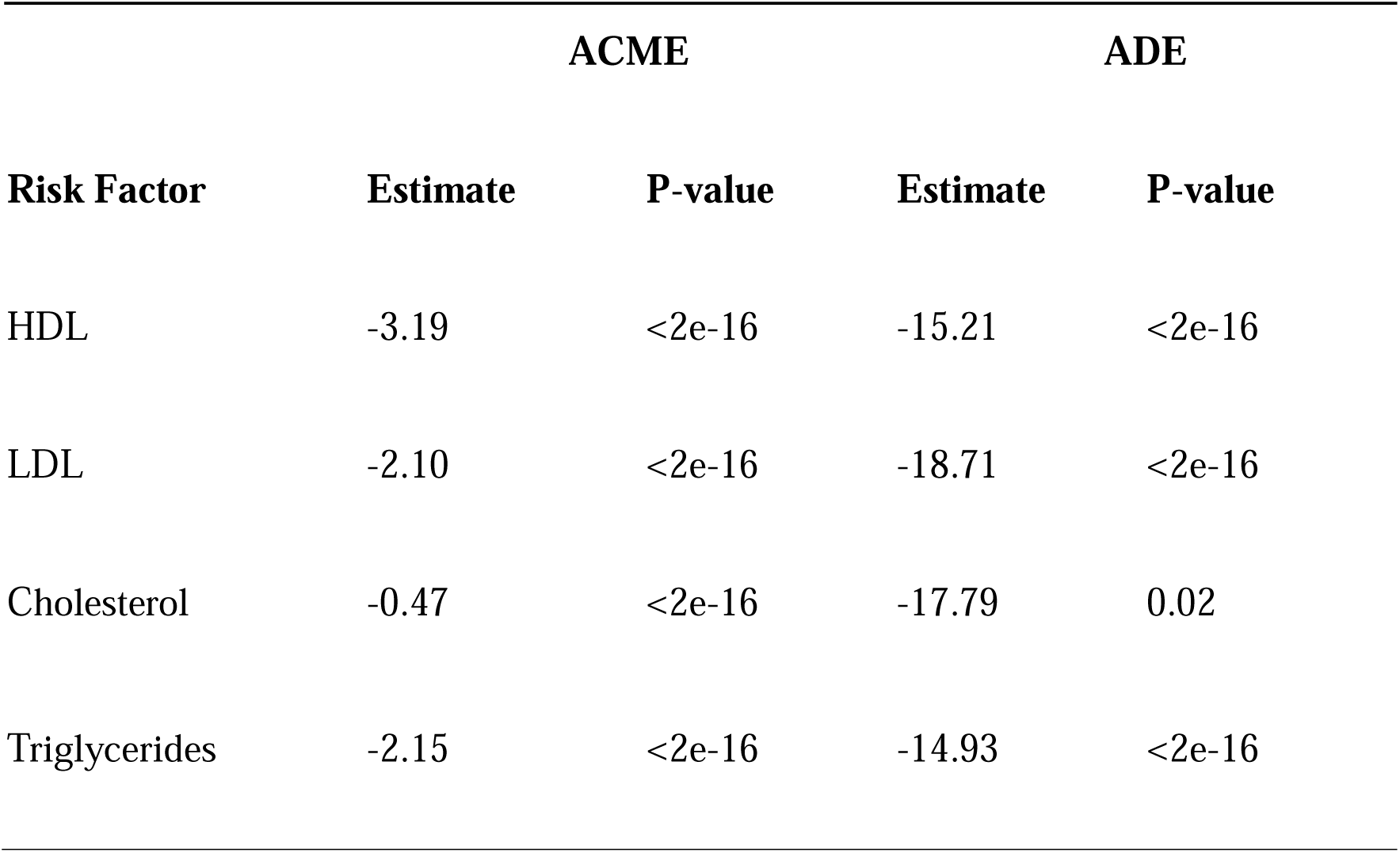
Results from the mediation analysis quantifying the effects select laboratory measurements have on the association between sleep disorders and incident aortic stenosis. Data from the TriNetX database**. ACME:** Average causal mediation effects of sleep disorders on aortic stenosis through the risk factor **ADE:** Average direct effect of sleep disorders on aortic stenosis

## Discussion

Using two separate databases, we report an increased risk for AS development in patients with a diagnosis of any sleep disorder. This statistically significant association was mainly driven by sleep apnea, hypersomnias, narcolepsy, and sleep movement disorders and remained robust when the model was controlled for traditional cardiovascular risk factors, suggesting that sleep disorders independently contribute to the AS risk profile. We further find that the increased risk is largely associated with a higher prevalence of classical cardiovascular risk factors and particularly dyslipidemia. We conclude that while dyslipidemia plays a role in mediating the increased risk for AS development, patients with sleep disorders are also at an increased risk for AS incidence through an independent mechanism. While sleep disorders were not attributed to the largest proportion of AS cases as compared to the classical cardiovascular risk factors, they are still an important modifiable target in cardio-protection. A population-based study reported 85% of incident AS cases required hospitalization for severe AS which underscores the need for preventive care targeting modifiable risk factors.^17^

Circadian rhythms are present in all physiologic and behavioral functions, amongst the most prominent of which is the sleep-wake cycle.^18^ These rhythms are the result of endogenous molecular clocks that are self-sustained transcriptional and translational feedback loops.^19^ Molecular clocks are found in most cells and tissues and influence cellular function by regulating transcription of large sets of genes.^20^ While being self-sustained, peripheral clocks are synchronized by a central master clock in the suprachiasmatic nucleus via hormonal or neural cues.^20^ Similar to other peripheral tissues, the molecular clock machinery is present in the heart and the vasculature and is synchronized with the clocks of other tissues by hormonal, neural, and food signals.^21^ As a consequence, cardiovascular function varies rhythmically across the 24 hour day ranging from electrophysiology, to blood pressure, to contractility, metabolic states, and remodeling and injury responses.^21–24^ In light of our results implicating lipid markers, it is important to note that lipid metabolism is under circadian control and varies rhythmically throughout the day.^25^ Knocking out components of the molecular clock in animal models has led to increased atherosclerotic burden and the development of different cardiomyopathies.^21, 26^ However, a potential impact of clocks on valvular physiology and disease has largely remained unaddressed.

Sleep apnea emerged with the largest effect size in our analysis of seven specific sleep disorder phenotypes. We speculate that a multiplicity of factors could be relevant to this observation. Obesity is an important risk factor for sleep apnea and is accompanied by metabolic dysfunction both of which are aortic stenosis risk factors.^9, 27^ The periods of breathlessness in sleep apnea increase sympathetic tone at night via chemoreflex stimulation by hypoxia, leading to both hypertension and metabolic dysfunction.^27^ A third factor is the disruption of normal circadian rhythms by apneic episodes and sympathetic activation. For hypersomnia and sleep movement disorders, evidence also suggests the involvement of circadian misalignment and dysautonomia.^28^

Genetic disruption of clock genes in rodents lead to various metabolic consequences such as increased body weight, hyperglycemia, hyperinsulinemia, and hyperlipidemia.^29^ Epidemiological studies have linked inadequate sleep to diabetes,^30^ hypertension,^31^ obesity,^32^ and hyperlipidemia.^33^ In a study from the UK biobank, sleep apnea was associated with coronary heart disease, and metabolic risk factors (especially dyslipidemia) played a mediating role.^34^ We see a similar effect where patients with sleep disorders had a higher prevalence of cardiometabolic risk factors and were at a higher risk for developing aortic stenosis.

Our study leverages large sample sizes of real-world EHR data from diverse populations across the US. The large sample size allowed a longitudinal design which provides confidence in the validity of observed associations. While we took measures to reduce the risk of reverse causation, it is not eliminated. Further studies with designs permitting causal conclusions are needed. This study also relies on EHR data, which by its nature, targets a sicker population than the general population. This could have diluted some signals leading us to miss associations that might have had relevance in the general population. Another potential limitation is the possibility of an overlap in participating patients since both databases are large cohort studies based in the US.

Patients with sleep disorders are at a higher risk of developing AS, an association largely mediated by dyslipidemia. Our study also suggests the presence of a distinct mechanism linking sleep disorders to the development of AS that is independent from the traditional cardiovascular risk factors including dyslipidemia. The application of machine learning to refine and integrate sleep and image derived phenotypes with genetics will elucidate the mechanisms which underlie this apparent relationship. Thus, sleep disorders are potential modifiable risk factors for the development of AS.

## Data Availability

All data used in this study are available by applying for access from the TriNetX network and the All of Us study.

## Acknowledgements

We gratefully acknowledge All of Us participants for their contributions, without whom this research would not have been possible. We also thank the National Institutes of Health’s All of Us Research Program for making available the participant data examined in this study.

## Supplementary Tables

**Table S1.** Codes used to extract data from both the TriNetX and All *of* Us data.

| Condition/Measurement | Code System | Codes |
| --- | --- | --- |
| Non-Rheumatic Aortic Stenosis | ICD-10 | I35.0, I35.2 |
| Non-Rheumatic Aortic Valve Disease | ICD-9 | 424.1 |
| Insomnia | ICD-9/ICD-10 | 327.0, 307.41, 307.42, G47.0, F51.0 |
| Hypersomnia | ICD-9/ICD-10 | 327.1, 307.43, 307.44, G47.1, F51.1 |
| Circadian Rhythm Sleep Disorder | ICD-9/ICD-10 | 327.3, 307.45, G47.2 |
| Sleep Apnea | ICD-9/ICD-10 | 327.2, G47.3 |
| Narcolepsy | ICD-9/ICD-10 | 347.0, G47.4, 307.4 |
| Parasomnia | ICD-9/ICD-10 | 327.4, 307.46, G47.5, F51.3, F51.4, F51.5 |
| Sleep Movement Disorders | ICD-9/ICD-10 | 327.5, G47.6 |
| Congenital Aortic Valve Disorders | ICD-9/ICD-10 | 746.3, 746.4, Q23.0, Q23.1 |
| Hypertension | ICD-9/ICD-10 | 401, 405, I10, I15 |
| Diabetes | ICD-9/ICD-10 | 249, 250, E08, E09, E10, E13 |
| Hyperlipidemia | ICD-9/ICD-10 | 272.0, 272.1, 272.2, 272.3, 272.4, E78.0, E78.1, E78.2, E78.3, E78.4, E78.5 |
| Chronic Kidney Disease | ICD-9/ICD-10 | 585, N18 |
| Aortic Valve Procedure | ICD-10/CPT/HCPCS | 1006141, 1006142, 1006147, 1006151, 1021150, 1029692, 1029693, 1035167, 33361, 33362, 33363, 33364, 33365, 33366, 33367, 33368, 33369, 33390, 33391, 33400, 333401, 333403, 333405, 333406, 333410, 333411, 333412, 333413, 33440, 33361, 92986, 939591, 027F04Z, 027F04Z, 027F0ZZ, 027F34Z, 027F3DZ, 027F3ZZ, 027F44Z, 027F4DZ, 027F4ZZ, 02QF, 02QF0ZJ, 02QF0ZZ, 02QF3ZJ, 02QF3ZZ, 02QF4ZJ, 02QF4ZZ, 02RF, 02RF07Z, 02RF08N, 02RF08Z, 02RF0JZ, 02RF0KZ, 02RF37H, 02RF37Z, 02RF38H, 02RF38N, 02RF38Z, 02RF3JH, 02RF3JZ, 02RF3KH, 02RF3KZ, 02RF47Z, 02RF48N, 02RF48Z, 02RF4JZ, 02RF4KZ, 02WF, 02WF07Z, 02WF08Z, 02WF0JZ, 02WF0KZ, 02WF37Z, 02WF38Z, 02WF3JZ, 02WF3KZ, 02WF47Z, 02WF48Z, 02WF4JZ, 02WF4KZ, X2RF, X2RF032, X2RF332, X2RF432, 35.01, 35.11, 35.21, 35.22 |
| Statin Use | RxNorm | 83367, 301542, 41127, 42463, 596723, 6472, 36567 |

**Table S2.** Prevalence of sleep disorders before age 65 and the mean age at diagnosis in the studied population from the TriNetX Network.

| **Sleep Disorder** | **Number**<br>**Diagnosed**<br>**before Age**<br>**65** | **Percent**<br>**Diagnosed**<br>**Before Age**<br>**65** | **Mean**<br>**Age at**<br>**Diagnosis**<br>**(± SD)** |
| --- | --- | --- | --- |
| All Sleep Disorders | 772085 | 10.58 | 62.74<br>($\pm 2.4$) |
| Sleep Apnea | 517569 | 7.09 | 62.76<br>($\pm 2.2$) |
| Circadian Rhythm Sleep Disorder | 6497 | 0.09 | 62.93<br>($\pm 2.3$) |
| Insomnia | 274981 | 3.77 | 62.9<br>($\pm 2.4$) |
| Hypersomnia | 35288 | 0.48 | 63.21<br>($\pm 2.1$) |
| Parasomnia | 6261 | 0.09 | 63.25<br>($\pm 2$) |
| Narcolepsy | 5937 | 0.08 | 62.28<br>($\pm 3.2$) |
| Sleep Movement Disorders | 29407 | 0.40 | 63.08<br>($\pm 2$) |

**Table S3.** Characteristics of the two patient populations in the All *of* US study from data collected before age 65.

| **Variable** | **No Sleep Disorder before**<br>**65 n (%)** | **Any Sleep Disorder before**<br>**65 n (%)** | **p** |
| --- | --- | --- | --- |
| n | 79296 | 7,839 |  |
| Hypertension | 17,101 (21.6) | 5,714 (72.9) | <0.001 |
| Diabetes | 6,670 (8.4) | 2,970 (37.9) | <0.001 |
| Hyperlipidemia | 17,662 (22.3) | 5,910 (75.4) | <0.001 |
| CKD | 1,835 (2.3) | 1,078 (13.8) | <0.001 |
| Sex (M) | 35,657 (45.0) | 3,495 (44.6) | 0.524 |
| BMI mean(sd) | 29.53 (6.01) | 33.94 (7.41) | <0.001 |

**Table S4.** Prevalence of sleep disorders before age 65 and the mean age at diagnosis in the studied population from the All *of* US study.

| **Sleep Disorder** | **Number**<br>**Diagnosed**<br>**before Age**<br>**65** | **Percent**<br>**Diagnosed**<br>**Before Age**<br>**65** | **Mean**<br>**Age at**<br>**Diagnosis**<br>**(± SD)** |
| --- | --- | --- | --- |
| All Sleep Disorders | 7839 | 9.00 | 60.67<br>± 3.9 |
| Sleep Apnea | 5628 | 6.46 | 60.78<br>± 3.5 |
| Circadian Rhythm Sleep Disorder | 180 | 0.21 | 60.71<br>± 3.9 |
| Insomnia | 2519 | 2.89 | 61.52<br>± 3.7 |
| Hypersomnia | 685 | 0.79 | 63.21<br>± 3.9 |
| Parasomnia | 109 | 0.13 | 61.37<br>± 3.6 |
| Narcolepsy | 82 | 0.09 | 57.33<br>± 6 |
| Sleep Movement Disorders | 488 | 0.56 | 61.1<br>± 3.1 |

**Table S5.** Regression model results for all labs tested including regression coefficients, *p-*values, Benjamini-Hochberg adjusted *q-*values, number of patients, and fold changes. Rare lab tests performed in <100 patients were analyzed but not included in the table to ensure anonymity. **(Table included as separate excel file.)**

**Table S6.** Means and standard deviation of each laboratory test result in each cohort for the lab tests with a *q-*value ≤0.1 and fold change ≥0.5 in all three cohort comparisons with the prior sleep disorder and incident AS cohort.

| Lab | cohort | mean | sd |
| --- | --- | --- | --- |
| Cholesterol | AS incidence and prior sleep disorder | 164.11 | 37.48 |
| Cholesterol | AS incidence with no prior sleep disorder | 178.95 | 39.63 |
| Cholesterol | No AS incidence with prior sleep disorder | 178.08 | 41.27 |
| Cholesterol | Healthy | 180.17 | 43.19 |
| HDL | AS incidence and prior sleep disorder | 46.84 | 13.43 |
| HDL | AS incidence with no prior sleep disorder | 49.13 | 14.23 |
| HDL | No AS incidence with prior sleep disorder | 51.22 | 14.54 |
| HDL | Healthy | 53.46 | 15.12 |
| LDL | AS incidence and prior sleep disorder | 93.94 | 29.68 |
| LDL | AS incidence with no prior sleep disorder | 103.15 | 31.26 |
| LDL | No AS incidence with prior sleep disorder | 103.04 | 30.45 |
| LDL | Healthy | 107.63 | 30.73 |
| Triglyceride | AS incidence and prior sleep disorder | 143.68 | 59.70 |
| Triglyceride | AS incidence with no prior sleep disorder | 134.27 | 58.99 |
| Triglyceride | No AS incidence with prior sleep disorder | 131.74 | 56.61 |
| Triglyceride | Healthy | 122.68 | 53.59 |

## Supplementary Figures

**FigureS1:**
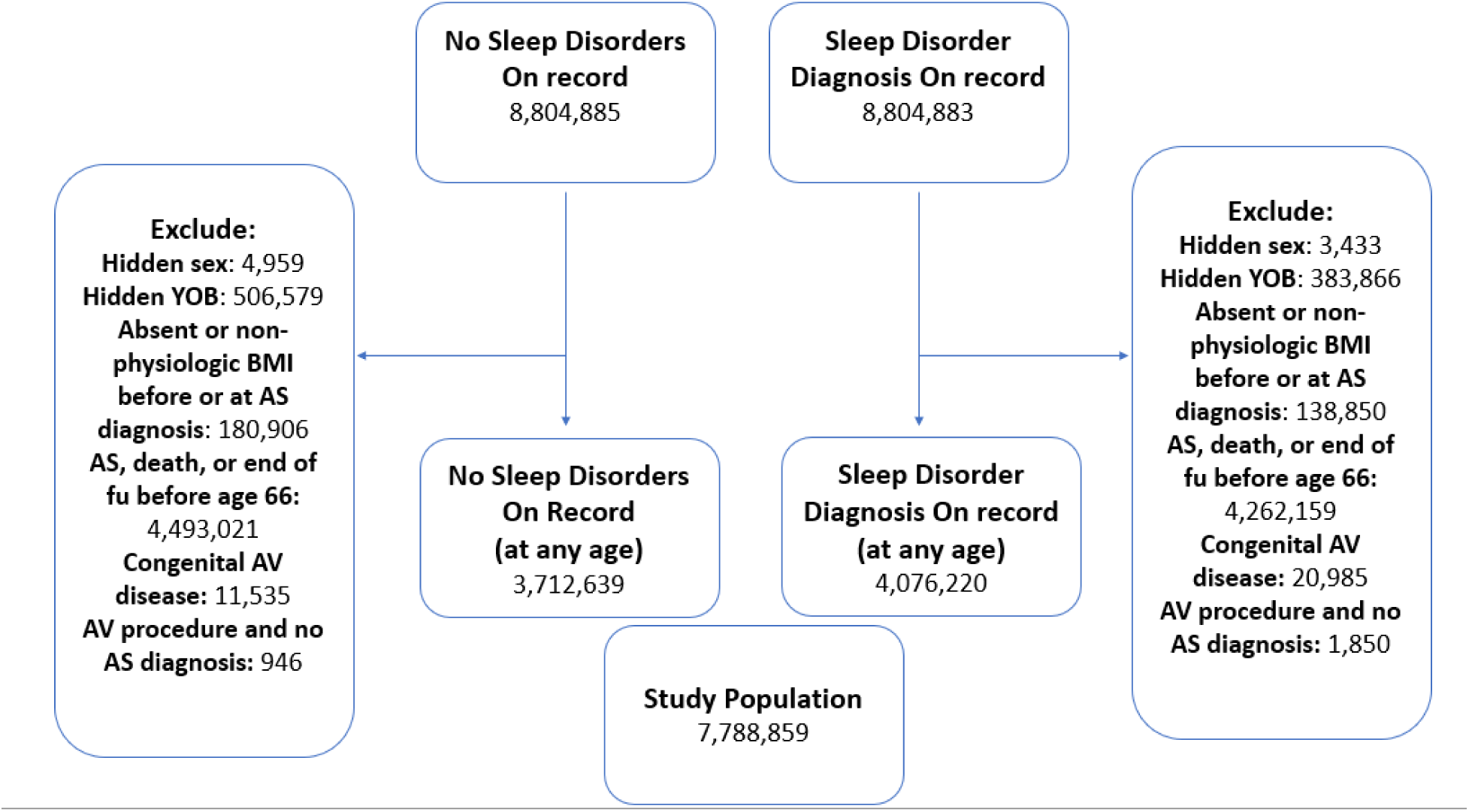
Flow chart representing the selection of patient records for inclusion in our analysis from the TriNetX network. YOB: year of birth, AS: aortic stenosis, fu: follow up, AV: aortic valve.

**FigureS2:**
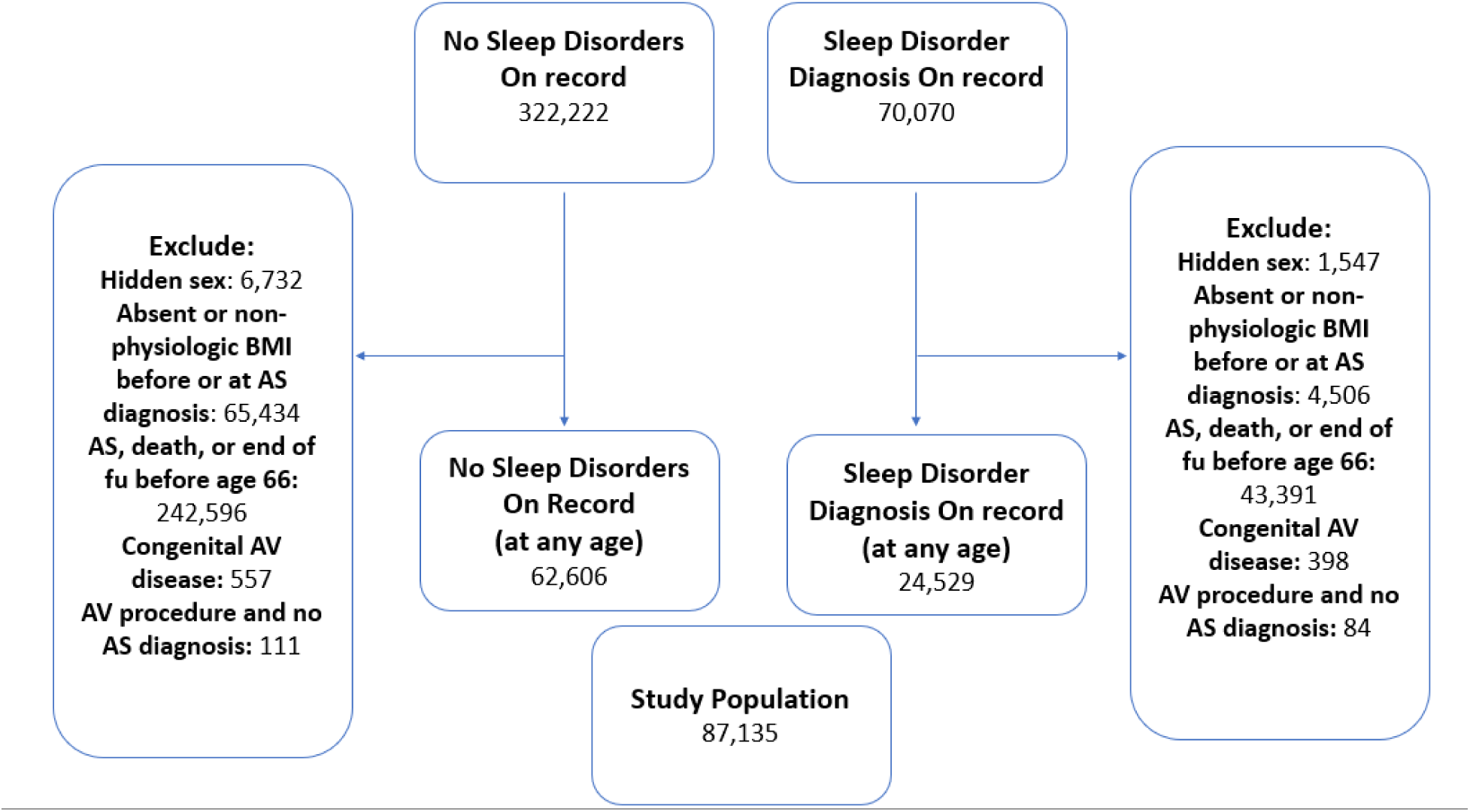
Flow chart representing the selection of patient records for inclusion in our analysis from the All *of* US study. YOB: year of birth, AS: aortic stenosis, fu: follow up, AV: aortic valve.

**Figure S3.**
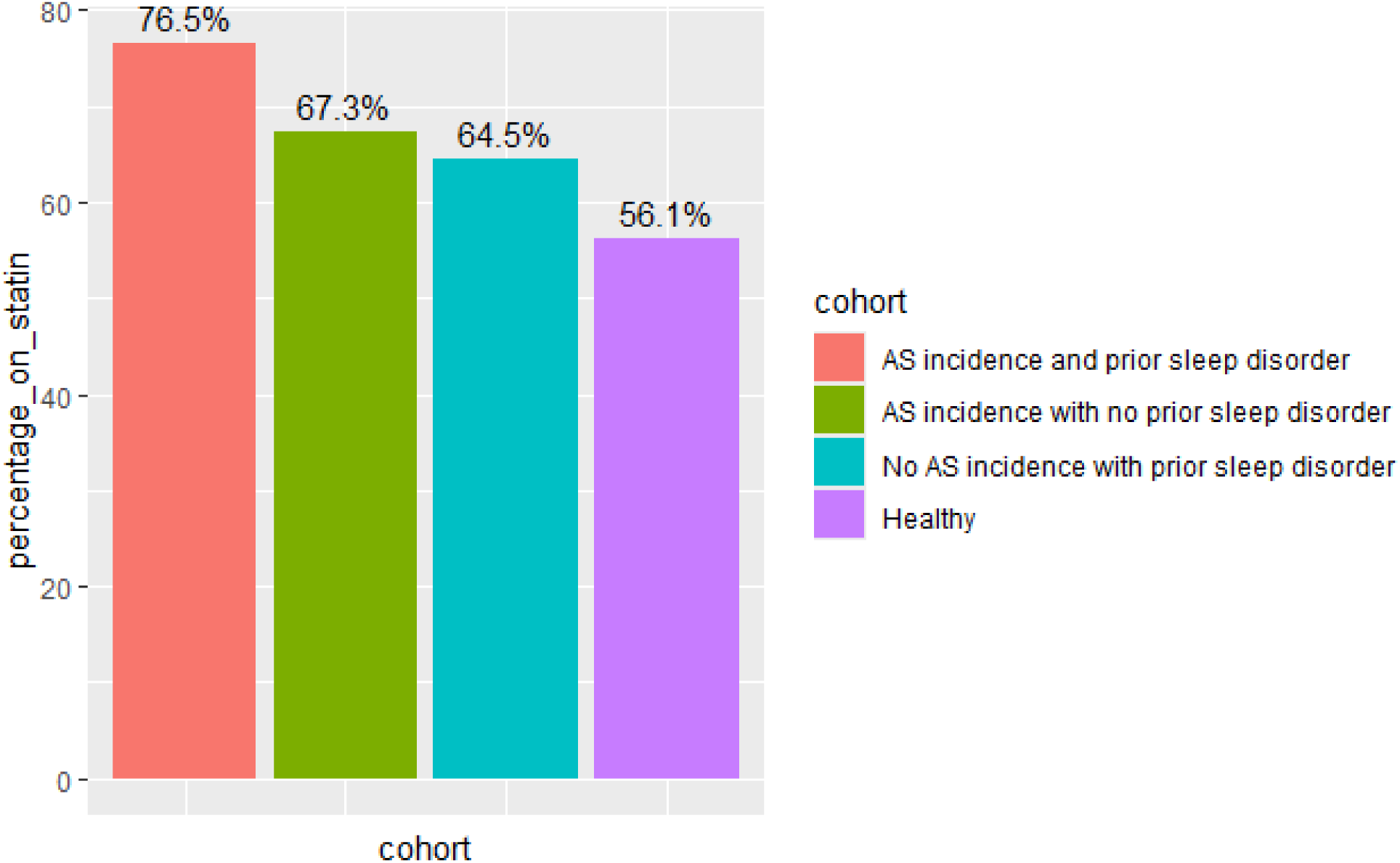
Percentage of patients prescribed any statin before they reached 65 years of age in each of the cohorts used in the laboratory test analysis. Chi-squared *p* value <0.001. Data from the TriNetX network.

